# Exploring the Factors Influencing Resilience Among Returnee Migrants in Nigeria

**DOI:** 10.64898/2026.07.02.26357141

**Authors:** Olatunji Joshua Awoleye, Olaniyi Felix Sanni, Kazeem Abimbola Uthman, Florence Ngozi Uchendu

## Abstract

**Background:** Returnee migrants in Nigeria often face significant psychosocial and economic challenges during reintegration, necessitating resilience to adapt and recover. This study examined factors influencing resilience among returnee migrants in Nigeria.

**Methodology:** A mixed-methods design was employed, involving 1,316 returnees selected through multistage sampling across Nigeria’s six geopolitical zones. Quantitative data were collected using the Connor–Davidson Resilience Scale (CD-RISC-25) and analyzed using SPSS version 28. Qualitative data was obtained through eight focus group discussions and analyzed thematically.

**Result:** Social support from family and friends was inconsistent (70.8% reported occasional support), while community support was largely absent (85.9%). Financial insecurity was widespread (>90%). Male gender (AOR = 6.092, p<0.001), ethnicity, and higher education were significant predictors of resilience. Qualitative findings highlighted the role of family support, faith, adaptive coping, and skill acquisition in strengthening resilience.

**Conclusion:** Resilience among returnee migrants in Nigeria is limited by weak structural and economic support, despite moderate personal coping capacity. Strengthening economic opportunities, community integration, and access to mental health services is essential for sustainable reintegration.

## Background

Returnee migrants often face complex challenges as they reintegrate into their home countries, including social and cultural adjustment, economic instability, and psychological distress. Understanding the factors that enhance resilience - defined as the ability to adapt positively in the face of adversity is crucial for supporting successful reintegration outcomes.

At the individual level, future orientation and hope are central to resilience. A forward-looking mindset and clearly defined goals empower returnees to overcome barriers and adapt to new realities [1]. This sense of optimism fosters psychological stability and motivation even amid economic uncertainty or social rejection. Similarly, religion and spirituality play vital roles in promoting emotional well-being and providing meaning during reintegration [2, 3].

Social support networks are equally critical for reintegration success. Family, friends, and peers provide emotional encouragement, financial aid, and practical assistance, all of which mitigate stress and facilitate adjustment [4]. Moreover, community engagement, through volunteering, cultural participation, or local governance, helps returnees rebuild social ties, regain purpose, and reduce isolation. Evidence indicates that active community involvement improves life satisfaction and strengthens social integration [5, 6]. Returnees who reconnect with their cultural roots tend to navigate adjustment more smoothly [7–9].

At the institutional level, access to education and employment opportunities serves as a foundation for economic independence and self-worth. Stable income and vocational training significantly enhance reintegration outcomes [10, 11]. Additionally, supportive policies and services, including legal aid, housing, and mental health support, are crucial for addressing structural barriers and promoting well-being [12, 13].

The reintegration of Nigerian returnee migrants remains a complex challenge involving economic, social, and psychosocial needs. According to the International Organization for Migration (2017), inadequate support from key stakeholders, including the IOM and the Nigerian government, under the Assisted Voluntary Return and Reintegration (AVRR) framework, has weakened reintegration efforts. Limited funding, difficulties in resource mobilization, and donor-related constraints have further reduced the effectiveness and sustainability of support for returnees [14].

Addressing these challenges requires a holistic approach that not only strengthens economic reintegration but also enhances returnees’ psychosocial resilience. Despite growing evidence, there is limited empirical data on resilience among returnee migrants in Nigeria. Therefore, this study aimed to assess the factors influencing resilience among returnee migrants in Nigeria.

## Methodology

### Research Design

This study utilised a mixed-method approach, combining quantitative and qualitative methods (Focus group discussion) to comprehensively assess the resilience factors among returnee migrants in Nigeria. This approach aligns with the overall research design, enabling a thorough investigation of the multifaceted issues.

### Study setting and Population

The research was conducted in Nigeria, West Africa’s most populous country noted for its cultural richness. Nigeria is divided into six geopolitical zones (Northwest, Northcentral, Northeast, Southwest, South-south, and Southeast), each with its own set of characteristics and challenges [15]. The research population consisted of returnee migrants in Nigeria.

### Inclusion criteria

1. Participants comprised individuals who have returned to Nigeria after a period of migration, whether voluntarily or forcibly, and have psychosocial needs.
2. Participants comprised individuals who can provide mature perspectives on their experiences and well-being.
3. Participants who willingly agreed to participate in the study and provide informed consent, ensuring their voluntary involvement.

### Exclusion Criteria

1. Participants who have not experienced migration and return to Nigeria.
2. Participants who cannot provide informed consent due to cognitive impairments, mental health issues, or any other factors that prevent them from fully understanding the nature of the study and voluntarily participating were excluded.

### Sample Size Determination

For the quantitative part, the sample size was determined using Yamane’s formula for a known population of 30,574 with a 5% margin of error, yielding an initial sample size of 395. This was adjusted for a design effect of 3 (to account for cluster sampling), increasing the sample to 1,185. After adding a 10% allowance for non-response, the final sample size was 1,316 respondents.

The qualitative phase involved 8 focus group discussions (FGDs) consisting of 12 participants each, broad representation and diversity in experience were ensured.

### Sampling Technique

A four-stage multistage sampling technique was employed. In the first stage, returnee migrants were stratified by Nigeria’s six geopolitical zones using available data from national and international migration databases. In the second stage, the number of participants selected from each zone was determined using probability proportional to size (PPS) to ensure representativeness. In the third stage, states within each zone were selected using the same PPS approach to maintain proportional allocation. In the final stage, convenience and snowball sampling techniques were used to recruit eligible participants, facilitating access to dispersed and hard-to-reach returnee populations.

For the qualitative component, participants were purposively selected based on their experiences of reintegration challenges and mental health difficulties to ensure rich, relevant insights.

### Data Collection and Instrument

Quantitative data were collected using a pretested interviewer-administered questionnaire and the Connor-Davidson Resilience Scale (CD-RISC 25) to assess resilience among returnee migrants in Nigeria [16]. The questionnaire was piloted and refined to improve clarity and respondent comfort.

Qualitative data were obtained through eight Focus Group Discussions (FGDs) conducted across geopolitical zones, each involving 12 participants. Discussions explored lived experiences, coping strategies, reintegration challenges, and cultural influences on mental health. All sessions were audio-recorded, transcribed verbatim, and analyzed thematically. Combining quantitative and qualitative findings strengthened the depth, reliability, and validity of the study.

### Validity Test and Reliability Test

Validity in this study was ensured through expert review to confirm content accuracy and comprehensive coverage of the resilience factors of returnee migrants. Criterion validity was further established by comparing the instrument with similar validated tools to confirm its predictive capability. Reliability was demonstrated through strong internal consistency (Cronbach’s Alpha = 0.80) and test-retest reliability, with a high correlation (r = 0.85) between responses collected at different times, confirming the instrument’s stability and consistency in measuring the intended constructs.

### Data Analysis

Data were initially entered and cleaned up in Excel, with about 10% of questionnaires discarded due to incomplete or inconsistent responses. The cleaned dataset was analyzed using IBM-SPSS version 28.0, employing descriptive such as frequency, and percentage was used to analyses to assess resilience factors of returnee migrants. For inferential analysis, for logistic regression. Bivariate analysis was first conducted using crude odds ratios (COR) to assess the association between independent variables (age, sex, religion, ethnicity, and education) and resilience. Variables with p-values <0.05 at the bivariate level were included in the multivariate logistic regression model to control for potential confounders. Adjusted odds ratios (AOR) with 95% confidence intervals (CI) were computed, and statistical significance was set at p < 0.05. For the qualitative component, audio recordings from FGD were transcribed verbatim and analyzed thematically. The analysis involved repeated reading, coding, and theme development to capture key patterns related to returnee migrants’ mental health and resilience. QDA Miner software supported data organization and management, providing deeper insights into the underlying reasons and processes shaping the experiences of returnee migrants and complementing the quantitative findings.

### Ethical Considerations

Ethical approval was sought and obtained from the Nigeria Institute of Medical Research (NIMR). The reference number for the ethical approval is (IRB/24/017). The Informed consent was obtained from all study participants, ensuring their voluntary and informed participation in the research. To protect participants’ privacy, measures were taken to guarantee anonymity and confidentiality regarding their responses and personal information. Ethical approval was also sought from NIMR to ensure that the study adheres to ethical standards and safeguards the well-being and rights of all participants.

## Results

### Socio-demographic Characteristics of Respondents

A total of 1,316 returnee migrants participated in the study. Most respondents were young adults aged 18–30 years (65.7%), with a mean age of 29 ± 6.2 years, while 33.5% were aged 31–50 years and only 0.5% were 51 years and above. Female respondents (56.5%) slightly outnumbered males (43.5%). More than half were single (58.3%), followed by married participants (38.4%), with few being divorced (1.8%), widowed (1.1%), or separated (0.4%). Regarding religion, most participants were Christians (79.0%), while 21.0% were Muslims. Ethnically, the largest group was Edo (50.9%), followed by Hausa (15.8%), Yoruba (9.7%), Igbo (7.9%), and other ethnic groups (15.7%). (Table 1).

**Table 1:** Socio-Demographic Characteristics of Returnee Migrants.

| Variable | Frequency (n) | Percentage (%) |
| --- | --- | --- |
| **Age (years)** |  |  |
| 0–17 | 5 | 0.4 |
| 18–30 | 864 | 65.7 |
| 31–50 | 441 | 33.5 |
| ≥51 | 6 | 0.5 |
| Mean age | 29 ± 6.2 |  |
| **Sex** |  |  |
| Male | 572 | 43.5 |
| Female | 744 | 56.5 |
| **Marital Status** |  |  |
| Single | 776 | 58.3 |
| Married | 506 | 38.4 |
| Separated | 5 | 0.4 |
| Divorced | 24 | 1.8 |
| Widowed | 14 | 1.1 |
| **Religion** |  |  |
| Christianity | 1040 | 79.0 |
| Islam | 276 | 21.0 |
| **Ethnicity** |  |  |
| Hausa | 208 | 15.8 |
| Igbo | 104 | 7.9 |
| Yoruba | 127 | 9.7 |
| Edo | 670 | 50.9 |
| Others | 207 | 15.7 |

### Assessment of Resilience Factors Among Returnee Migrants

Table 2 shows that most respondents reported moderate but inconsistent access to support systems and coping mechanisms. Social support from family and friends was relatively strong, with the majority (70.8%) indicating it was *sometimes true*, though very few reported frequent support. In contrast, community support was generally weak, with most respondents (69.3%) stating it was *rarely true*. Financial stability was notably low, as a large proportion (59.9%) reported it was *not true at all*, and 31.3% said it was *rarely true*. Similarly, participation in resilience activities was limited, with most respondents indicating it was *rarely true* (57.1%). For personal coping, most respondents reported *rarely* (61.9%) or *sometimes* (34.7%) engaging in coping strategies. Psychological support and emotional well-being strategies were more moderately available, with responses largely clustered around *rarely* and *sometimes true*.

**Table 2:** Assessment of Resilience factors of respondents among Returnees migrants.

| Response | Frequency (n) | Percentage (%) |
| --- | --- | --- |
| **Social support from family and friends** |  |  |
| Not true at all | 49 | 3.7 |
| Rarely true | 317 | 24.1 |
| Sometimes true | 932 | 70.8 |
| Often true | 6 | 0.5 |
| Nearly always true | 12 | 0.9 |
| **Community support** |  |  |
| Not true at all | 218 | 16.6 |
| Rarely true | 912 | 69.3 |
| Sometimes true | 30 | 2.3 |
| Often true | 62 | 4.7 |
| Nearly always true | 94 | 7.1 |
| **Financial stability** |  |  |
| Not true at all | 788 | 59.9 |
| Rarely true | 412 | 31.3 |
| Sometimes true | 14 | 1.1 |
| Often true | 95 | 7.2 |
| Nearly always true | 7 | 0.5 |
| **Resilience activities** |  |  |
| Not true at all | 362 | 27.5 |
| Rarely true | 752 | 57.1 |
| Sometimes true | 188 | 14.3 |
| Often true | 7 | 0.5 |
| Nearly always true | 7 | 0.5 |
| **Personal coping** |  |  |
| Not true at all | 28 | 2.1 |
| Rarely true | 814 | 61.9 |
| Sometimes true | 456 | 34.7 |
| Often true | 9 | 0.7 |
| Nearly always true | 9 | 0.7 |
| **Psychological support** |  |  |
| Not true at all | 153 | 11.6 |
| Rarely true | 562 | 42.7 |
| Sometimes true | 577 | 43.8 |
| Often true | 16 | 1.2 |
| Nearly always true | 8 | 0.6 |
| **Emotional well-being strategies** |  |  |
| Not true at all | 14 | 1.1 |
| Rarely true | 553 | 42.0 |
| Sometimes true | 726 | 55.2 |
| Often true | 14 | 1.1 |
| Nearly always true | 9 | 0.7 |

### Factors Associated with Resilience Levels

Table 3 shows that sex and ethnicity were the main significant predictors after adjustment. Males had significantly higher odds compared to females (AOR = 6.09, p < 0.001). For ethnicity, both Igbo (AOR = 0.45, p = 0.032) and Yoruba (AOR = 0.50, p = 0.039) had significantly lower odds compared to Hausa. Age, religion, and education were not statistically significant in the adjusted model, although tertiary education showed a borderline association (p = 0.055).

**Table 3:**
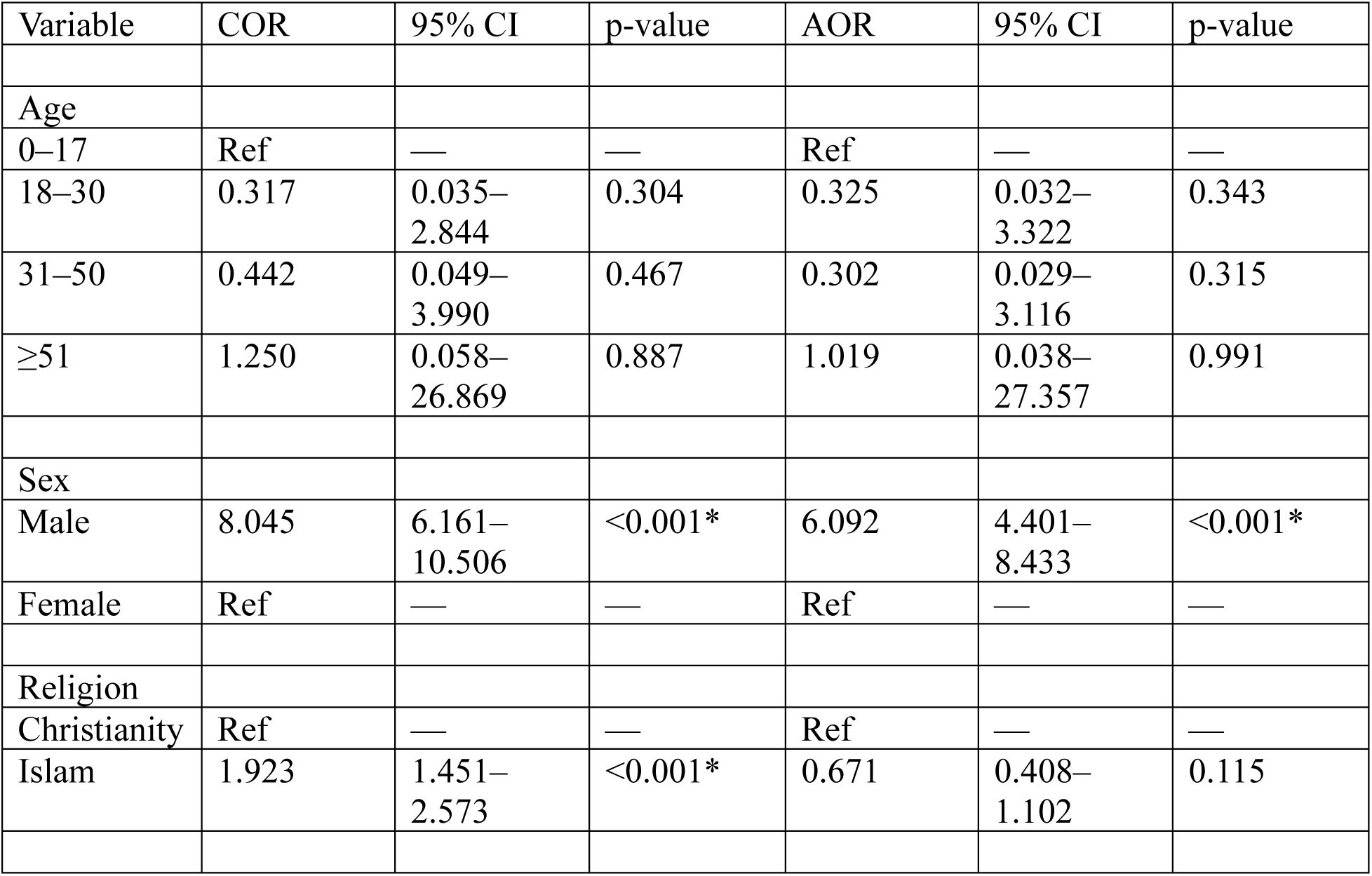
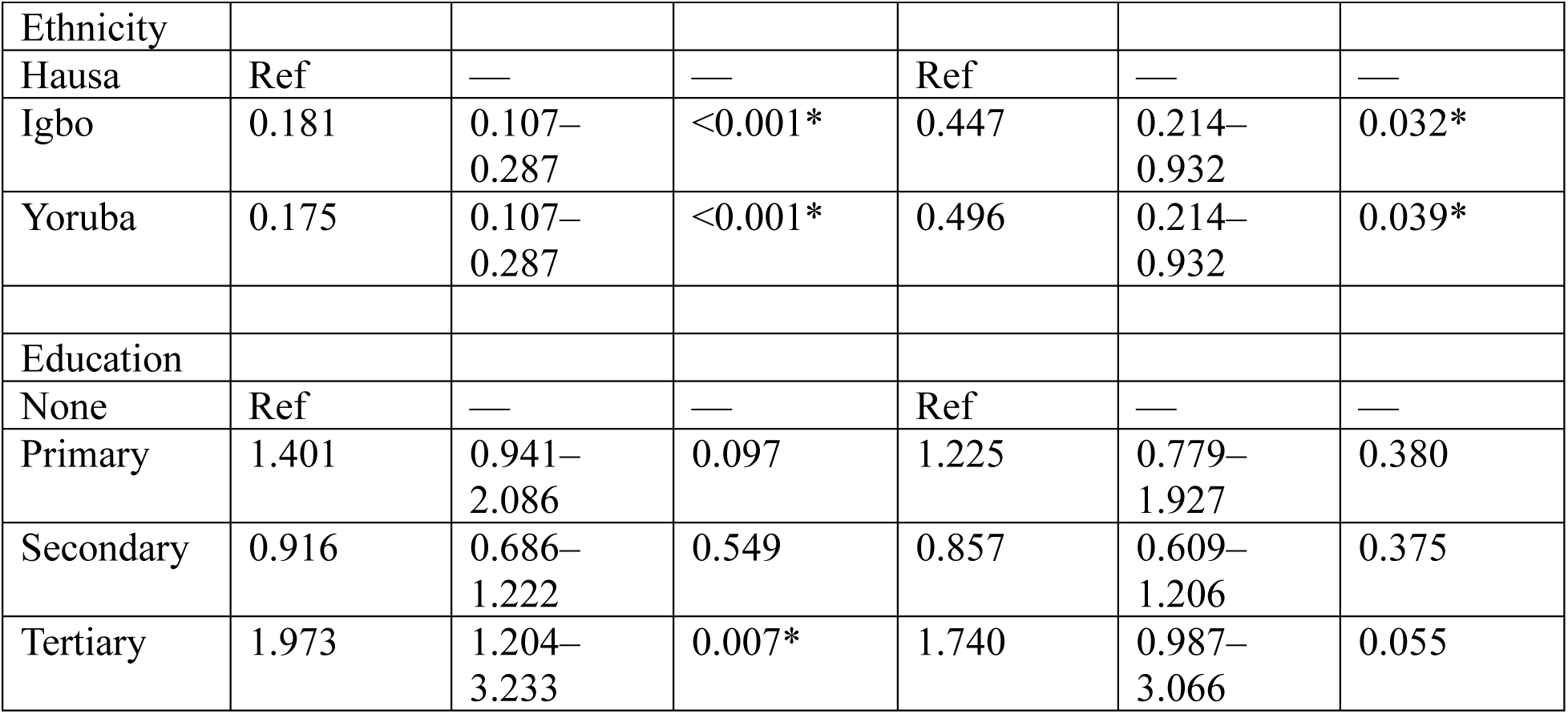
Multivariate Logistic Regression Analysis of Factors Associated with Resilience Levels among Returnee Migrants.

| Variable | COR | 95% CI | p-value | AOR | 95% CI | p-value |
| --- | --- | --- | --- | --- | --- | --- |
| Age |  |  |  |  |  |  |
| 0–17 | Ref | — | — | Ref | — | — |
| 18–30 | 0.317 | 0.035–<br>2.844 | 0.304 | 0.325 | 0.032–<br>3.322 | 0.343 |
| 31–50 | 0.442 | 0.049–<br>3.990 | 0.467 | 0.302 | 0.029–<br>3.116 | 0.315 |
| ≥51 | 1.250 | 0.058–<br>26.869 | 0.887 | 1.019 | 0.038–<br>27.357 | 0.991 |
| Sex |  |  |  |  |  |  |
| Male | 8.045 | 6.161–<br>10.506 | <0.001* | 6.092 | 4.401–<br>8.433 | <0.001* |
| Female | Ref | — | — | Ref | — | — |
| Religion |  |  |  |  |  |  |
| Christianity | Ref | — | — | Ref | — | — |
| Islam | 1.923 | 1.451–<br>2.573 | <0.001* | 0.671 | 0.408–<br>1.102 | 0.115 |
| Ethnicity |  |  |  |  |  |  |
| Hausa | Ref | — | — | Ref | — | — |
| Igbo | 0.181 | 0.107–<br>0.287 | <0.001* | 0.447 | 0.214–<br>0.932 | 0.032* |
| Yoruba | 0.175 | 0.107–<br>0.287 | <0.001* | 0.496 | 0.214–<br>0.932 | 0.039* |
| Education |  |  |  |  |  |  |
| None | Ref | — | — | Ref | — | — |
| Primary | 1.401 | 0.941–<br>2.086 | 0.097 | 1.225 | 0.779–<br>1.927 | 0.380 |
| Secondary | 0.916 | 0.686–<br>1.222 | 0.549 | 0.857 | 0.609–<br>1.206 | 0.375 |
| Tertiary | 1.973 | 1.204–<br>3.233 | 0.007* | 1.740 | 0.987–<br>3.066 | 0.055 |

### Qualitative Result

#### Coping Mechanisms and Strategies for Managing Reintegration Challenges

Returnee migrants adopt various strategies to manage the difficulties of reintegrating into Nigerian society. Their coping mechanisms reflect social and institutional support, and adaptive lifestyle changes.

#### Social Support Systems

Family, friends, and community networks were vital for emotional and financial recovery, though some experienced rejection.

“*My family showed me love and care*.” — R6, Ikeja FGD

“*My father shared his business with me*.” — R4, Ikeja Lagos FGD

“*I have a community of non-judgmental women who provide a support group*.” — R5, Ikorodu Lagos FGD

#### Environmental and Lifestyle Adjustments

Some returnees relocated, adopted new habits, or found comfort in creative and life-affirming activities.

“*I relocated from my community where I lived before traveling*.” — R1, Ikorodu Lagos FGD “*I create content and make TikTok videos*.” — R5, Ikeja Lagos FGD

“*Being pregnant and looking forward to meeting my baby comforted me*.” — R7, Ikeja Lagos FGD

Overall, returnee migrants combine personal resilience, social networks, and institutional assistance to navigate the challenges of reintegration.

### Personal Strengths and Community Resources Supporting Psychological Well-Being

Returnee migrants identified personal strengths and community supports that enhanced their psychological resilience during reintegration. Two main themes emerged: internal coping capacities and external support systems.

### Personal Strengths and Coping Mechanisms

Returnees demonstrated emotional control, purpose-driven engagement, and optimism in rebuilding their lives.

“*I cry when I feel overwhelmed, and afterwards I feel better*.” — R4, Ikeja Lagos FGD “*I love pampering myself and avoiding physical stress*.” — R5, Kano FGD

“*My skills keep me busy, so I don’t overthink*.” — R4, Ikorodu Lagos FGD

“*My resilient spirit keeps me going; I believe my life will get better*.” — R1, Ikorodu Lagos FGD

### Community Resources and Support Systems

Support systems ranged from institutional and religious aid to family networks and social interactions.

“*IOM supported me through theatre and art activities*.” — R5, Ikeja Lagos FGD “*My church welcomed me after my return*.” — R5, Ikeja Lagos FGD

“*My mom counsels and cheers me up*.” — R3, Ikorodu Lagos FGD

“*I attend football discussions and church programs to feel connected*.” — R4, Ikorodu Lagos FGD

### Some returnees, however, experienced community rejection or stigma

“*There is no support from the community*.” — R1, Ikorodu Lagos FGD “*The community has only stigmatized me*.” — R3, Kano FGD

Overall, returnees’ psychological well-being relies on both inner resilience (adaptability, hope, and purpose) and the strength of external networks (family, community, and institutional support).

### Impact of Relationships on Mental Health After Return

Interpersonal relationships strongly shaped returnees’ mental health, ranging from emotional support to rejection and stigma. Family, friends, and community dynamics either aided recovery or deepened distress. Three key themes emerged.

### Family Relationships: Support and Strain

Some families offered love and encouragement, while others caused emotional harm or exclusion. “*My mom shows me love and care*.” — R1, Ikorodu Lagos FGD

“*My father’s motivation and support helped me*.” — R4, Ikorodu Lagos FGD

“*I avoided my family due to stigma… my mother asked why I failed when others succeeded*.” —

R4, Kano FGD

“*My family did not know I returned*.” — R5, Kano FGD

### Friends and Peer Networks: Vital Support Systems

Friends, especially new ones and fellow returnees, provided crucial emotional and financial support.

“*A stranger turned friend supported me more than my old friends*.” — R6, Ikeja Lagos FGD “*I met a lady online who paid for my accommodation*.” — R8, Ikeja Lagos FGD

“*Only fellow returnees truly support one another*.” — R1, Ikorodu Lagos FGD

### Community Interactions: Acceptance and Stigma

Religious and social groups often offered belonging, but stigma and indifference were also common.

“*My church community helped me*.” — R3, Ikeja Lagos FGD

“*Negative impact through rejection and mockery*.” — R4, Ikeja Lagos FGD

“*My community treats me indifferently; everything is just normal*.” — R3, Abuja FGD

Overall, relationships were either a powerful source of healing or a trigger for emotional distress, depending on their quality and acceptance.

### Cultural and Religious Influences on Mental Resilience

Cultural and religious practices played mixed roles in shaping returnees’ mental resilience. While faith and spirituality offered emotional stability for some, others faced cultural pressure.

### Religion as a Source of Strength

Faith communities provided emotional grounding, hope, and social connection.

“*Hope and faith bring stability of belief… through meditation groups*.” — R5, Ikeja Lagos FGD “*My church embraced me and it helped*.” — R4, Ikorodu Lagos FGD

“*My religion reaches out to encourage me often*.” — R7, Ikorodu Lagos FGD

### Cultural Pressures and Expectations

Traditional norms around success and gender created emotional strain.

“*Cultural pressure places timelines on women to achieve things like marriage*.” — R1, Ikorodu Lagos FGD

“*They care more about what returnees bring back materially*.” — R1, Kano FGD “*Some cultures see traveling abroad as the only success path*.” — R3, Yobe FGD

Overall, religion often anchored emotional resilience, while cultural expectations and stigma sometimes intensified psychological stress, revealing both protective and harmful influences.

### Key Experiences and Resources Strengthening Resilience

Returnees identified multiple factors that supported their reintegration, reflecting a blend of institutional help, social connections, and personal growth.

### Institutional Support and Skills Development

Structured programs like those from IOM and GIZ provided critical economic stability and confidence.

“*The IOM vocational training on woodwork transformed my career prospects*.” — R5, Kano 2 FGD

“*Skill acquisition programs rebuilt my confidence*.” — R7, Ikeja FGD

### Community and Peer Networks

Social ties offered emotional and practical support, fostering belonging and shared understanding. “*Our returnee association became my second family*.” — R4, Benin City FGD

“*Fellow survivors from Libya stay connected and encourage me*.” — R2, Yobe 2 FGD

### Personal Growth and Inner Strength

Hardship inspired self-motivation, hope, and emotional healing.

“*Surviving Libya taught me endurance and determination*.” — R2, Lagos FGD “*Sharing my story empowers others and heals me*.” — R3, Ikorodu Lagos FGD

Resilience among returnees stemmed from a balance of external support, social connection, and personal transformation highlighting that recovery is sustained by both practical opportunities and inner strength.

## Discussion

This study revealed generally low levels of structural resilience among returnee migrants, particularly in relation to community support, financial stability, and access to mental health services. The assessment of resilience factors revealed generally low levels of protective mechanisms among returnee migrants. While 70.8% reported having some level of family and friends’ support, they indicated that this support was only sometimes true. This suggests that although social ties exist, they may not be sufficiently reliable or sustained to foster strong resilience. Community support, however, was particularly weak, with over two-thirds (69.3%) saying it was rarely true. This indicates a critical gap in collective or institutional reintegration structures. Community integration is a key factor influencing resilience, and its absence is associated with social exclusion and poorer mental health outcomes among returnees. Financial instability and low participation in recreational or faith-based activities were also commonly reported, indicating weak structural and personal resilience supports. This suggests that many returnees have limited access to adaptive coping mechanisms necessary for effective stress management and emotional well-being. These findings are consistent with those of Jayakody et al. (2022), who found that displaced populations experiencing poor community support and economic hardship demonstrated lower resilience. However, despite these challenges, many respondents still reported moderate personal resilience, relying on inner strength, hope, and determination, which aligns with studies showing that African migrants often depend on spirituality and optimism as coping strategies [17, 18].

Utilization of professional psychological support was also low, with the majority reporting “rarely” or “sometimes” seeking such services. This reflects known barriers in Nigeria, including stigma, cost, and limited access to mental health services. According to Srikanth et al. study Nigeria has a significant treatment gap, with fewer than 10% of individuals needing mental health care receiving it [19]

The regression analysis provides additional insights into predictors of resilience. Male respondents were significantly more likely to exhibit higher resilience compared to females (AOR = 6.092, p<0.001). This gender disparity may reflect socio-cultural norms in Nigeria that favor male economic participation and autonomy, which are critical for resilience. These support findings from Ghana and Ethiopia, where women migrants reported lower coping ability due to compounded risks of sexual exploitation, stigma, and caregiving burdens [20]. This finding should be interpreted cautiously due to potential residual confounding and sociocultural influences that the model may not fully capture.

Ethnicity also showed significant associations, with Igbo and Yoruba respondents having lower odds of resilience compared to Hausa respondents. This may reflect differences in migration experiences, social cohesion, or support systems for regional reintegration. However, the literature on ethnic disparities in migrant resilience in Nigeria is limited, underscoring the need for further research.

Education showed mixed effects, but higher educational attainment (tertiary level) was associated with increased resilience in the crude analysis and remained influential after adjustment. This aligns with existing evidence that education enhances problem-solving skills, access to opportunities, and adaptive capacity [21]. Age and religion were not significant predictors after adjustment, suggesting that resilience among returnees may be more strongly influenced by socio-economic and structural factors than demographic characteristics alone.

The findings from FGD participants indicate that returnees rely on a combination of economic empowerment, social support, and psychological coping strategies to adapt to life in Nigeria. Economic empowerment through skill utilization and vocational training was particularly effective, as participants often used pre-existing or newly acquired skills to reestablish livelihoods. This aligns with studies that emphasize the importance of sustainable income generation in reducing post-return vulnerability. For example, returnees who accessed vocational training or business support programs from IOM and GIZ gained not only financial stability but also a renewed sense of self-worth. These support findings from Akaeze & Shaibu research where deported immigrants in Nigeria considered potential for brain gain and economic growth if provided with proper support, such as comprehensive reintegration programs, vocational training, financial assistance, and social services [22].

Social support systems were another critical enabler. Strong family bonds, assistance from friends, and peer solidarity within returnee networks (like FREMNET) created protective environments. However, the findings also highlight the duality of these relationships; while some returnees benefited from love and material support, others experienced stigma, rejection, and indifference.

This study showed that successful reintegration largely depends on supportive family, social, and community relationships. The findings are consistent with Nte, study which reported that returnees benefited from community support, counselling services, educational assistance, economic empowerment, and family-based support systems [23]. Returnees used different coping strategies, including self-motivation and engagement in meaningful activities, although some adopted unhealthy behaviours such as alcohol use and dependence on sleeping tablets. Structured reintegration programmes and NGOs helped promote healthier coping mechanisms through mentorship, training, and community activities. Internal resilience factors such as emotional control, adaptability, hopefulness, and a sense of purpose also strengthened reintegration efforts. However, community support was not equally available to all participants, as some experienced stigma and lack of acceptance, highlighting the need for greater community sensitization and inclusive support for returnees.

These study findings also highlight that relationships emerged as a determinant of mental health outcomes for returnees. Families, when supportive, were the strongest buffers against distress, but when rejected or estranged, they were equally harmful. Friends and new acquaintances often stepped in as unexpected allies, with some offering financial or emotional lifelines. Peer networks, particularly returnee associations, created solidarity through shared experiences, thereby reducing isolation. This is in line with studies that showed that higher family support is associated with lower depressive symptoms and anxiety symptoms in adolescents [24, 25]. Conversely, stigmatizing community interactions perpetuated trauma and exclusion. The findings indicate the need for interventions that enhance family counseling, encourage peer support systems, and involve community leaders to reduce stigma and improve social relationships. Religion and culture were found to have both positive and negative influences on resilience. While faith provided hope, emotional support, and practical assistance for many returnees, some also experienced religious stigma and cultural pressure, particularly related to gender roles and expectations of financial success after migration. These findings emphasize the importance of culturally sensitive interventions that collaborate with faith-based and community institutions to promote supportive environments for returnees.

### Limitations and Strengths of the Study

This study has some limitations that should be acknowledged. The use of convenience and snowball sampling at the final stage may limit the generalizability of the findings. Furthermore, the reliance on self-reported data introduces the potential for recall and social desirability bias. Finally, the cross-sectional design precludes causal inference, and the observed associations should therefore be interpreted with caution.

Despite these limitations, this study has several strengths. It employed a mixed-methods design, which allowed for both quantitative measurement and in-depth qualitative exploration of resilience among returnee migrants. The relatively large sample size and inclusion of participants across Nigeria’s six geopolitical zones enhance the representativeness and breadth of the findings. Furthermore, the use of a validated instrument (CD-RISC-25) strengthens the reliability of the quantitative assessment. The integration of qualitative and quantitative data also enabled triangulation, providing a more comprehensive understanding of the factors influencing resilience

### Implication for Policy and Practice

Notwithstanding these limitations, the findings of this study offer practical implications for policy and practice. Efforts should be made to strengthen community-based support systems and reduce stigma to foster successful reintegration. Economic empowerment programs, such as vocational training, should be expanded to mitigate financial instability. Additionally, expanding access to culturally appropriate mental health services and fostering collaboration between governmental and community organizations will be vital in enhancing resilience outcomes for returnee migrants.

## Conclusion

This study demonstrates that returnee migrants in Nigeria face significant gaps in key resilience-supporting factors, particularly in community support, financial stability, and access to structured coping resources. Although some level of personal and social resilience exists, it remains insufficient to offset broader systemic challenges. Targeted interventions that integrate economic empowerment, community reintegration, and accessible mental health services are essential to strengthen resilience and promote sustainable reintegration outcomes among returnees. The findings underscore the importance of strengthening social support systems, promoting inclusive community acceptance, and expanding psychosocial and economic empowerment programs through collaborative government and non-governmental efforts. Enhancing these support structures will not only improve individual resilience but also facilitate sustainable reintegration and long-term well-being among returnee migrants.

## Data Availability

All data produced in the present study are available upon reasonable request to the authors
All data produced in the present work are contained in the manuscript

## Acknowledgment

The authors sincerely thank all returnee migrants who participated in this study, as well as the organizations and field staff who supported data collection across Nigeria. Their cooperation and contributions were essential to the successful completion of this research.

## Funding

This research received no external funding

## Conflict of Interest

There is no conflict of interest among the authors

## Notes

### Competing Interest Statement

The authors have declared no competing interest.

### Author Declarations

Ethical approval was sought and obtained from the Nigeria Institute of Medical Research (NIMR). The reference number for the ethical approval is (IRB/24/017).

